# MIXTURE of human expertise and deep learning—Developing an explainable model for predicting pathological diagnosis and survival in patients with interstitial lung disease

**DOI:** 10.1101/2021.07.21.21260920

**Authors:** Wataru Uegami, Andrey Bychkov, Mutsumi Ozasa, Kazuki Uehara, Kensuke Kataoka, Takeshi Johkoh, Yasuhiro Kondo, Hidenori Sakanashi, Junya Fukuoka

**Affiliations:** Department of Pathology, Nagasaki University Graduate School of Biomedical Sciences, Nagasaki, Japan; Department of Pathology, Kameda Medical Center, Kamogawa, Japan; Artificial Intelligence Research Center, National Institute of Advanced Industrial Science and Technology, Tsukuba, Ibaraki, Japan; Department of Respiratory Medicine and Allergy, Tosei General Hospital, Seto, Japan; Department of Radiology, Kinki Central Hospital of Mutual Aid Association of Public Health Teachers, Itami, Japan

**Keywords:** deep learning, artificial intelligence, explainable AI (xAI), machine learning, interstitial pneumonia, pulmonary fibrosis

## Abstract

Interstitial pneumonia is a heterogeneous disease with a progressive course and poor prognosis, at times even worse than those in the main cancer types. Histopathological examination is crucial for its diagnosis and estimation of prognosis. However, the evaluation strongly depends on the experience of pathologists, and the reproducibility of diagnosis is low.

Herein, we propose MIXTURE (huMan-In-the-loop eXplainable artificial intelligence Through the Use of REcurrent training), a method to develop deep learning models for extracting pathologically significant findings based on an expert pathologist’s perspective with a small annotation effort. The procedure of MIXTURE consists of three steps as follows. First, we created feature extractors for tiles from whole slide images using self-supervised learning. The similar looking tiles were clustered based on the output features and then pathologists integrated the pathologically synonymous clusters. Using the integrated clusters as labeled data, deep learning models to classify the tiles into pathological findings were created by transfer-learning the feature extractors. We developed three models for different magnifications.

Using these extracted findings, our model was able to predict the diagnosis of usual interstitial pneumonia, a finding suggestive of progressive disease, with high accuracy (AUC 0.90). This high accuracy could not be achieved without the integration of findings by pathologists. The patients predicted as UIP had significantly poorer prognosis (five-year overall survival [OS]: 55.4%) than those predicted as non-UIP (OS: 95.2%). The Cox proportional hazards model for each microscopic finding and prognosis pointed out dense fibrosis, fibroblastic foci, elastosis, and lymphocyte aggregation as independent risk factors. We suggest that MIXTURE may serve as a model approach to different diseases evaluated by medical imaging, including pathology and radiology, and be the prototype for artificial intelligence that can collaborate with humans.

## 1. Introduction

Interstitial pneumonia is a heterogenous benign disease that is subclassified based on histological features[1]. Idiopathic pulmonary fibrosis (IPF), for example, is a progressive condition with a 5-year survival probability of 45%[2], which is worse than that of major malignancies such as breast carcinoma, colorectal carcinoma, and cancers of the kidney and uterus[3]. It is treated with antifibrotic drugs to alleviate its progression[4, 5], and the treatments and outcomes are largely different from other types of interstitial pneumonia. Histologically, it is characterized by heterogeneously distributed destructive dense fibrosis predominating at the periphery and fibroblastic foci, which is known as the usual interstitial pneumonia (UIP) pattern[6].

Also in the interstitial pneumonia family, connective tissue disease-interstitial lung disease (CTD-ILD) represents one of the systemic manifestations of connective tissue disease, which include rheumatoid arthritis[7], Sjögren’s syndrome, systemic sclerosis[8], etc. It is known to have nonspecific interstitial pneumonia (NSIP) patterns as well as UIP patterns and is characterized by a variety of findings, including lymphoplasmacytic inflammation. Corticosteroids and immunosuppressive agents are commonly used for treatment[9]. Some other types of ILD, such as immune deficiency related interstitial pneumonia[10] and hypersensitivity pneumonia[11, 12] require different treatment protocols. In order to make an appropriate diagnosis, determine the prognosis and choose a therapeutic strategy, it is necessary for clinical, radiological, and pathological findings to be examined from multidisciplinary perspectives[13, 14], of which, pathological findings are particularly important[1]. However, it has been repeatedly pointed out that histological evaluation has a low concordance rate and reproducibility, which hinders the determination of treatment strategies and the understanding of pathogenesis[15–17].

Recent advances in whole slide imaging (WSI) and artificial intelligence (AI) technology, such as deep learning-based image processing, have opened the door to quantitatively evaluate histopathological findings[18]. Interestingly, WSI has added value in the pathological diagnosis of interstitial pneumonia because it allows easy observation of specimens on low-power magnifications (including those not available using a conventional microscope), which is important to recognize certain morphologic patterns with diagnostic significance[19].

The traditional pathological approach to diagnosis is to identify different microscopic findings, analyze the relationship between them, integrate data based on their professional experience, and eventually reach to the appropriate diagnosis regarded as a ground truth. Since pathology is critical for understanding pathogenesis and determining treatment strategies, recent reports have emphasized the importance of mechanisms that provide the explanation of the model’s outputs. Grad-CAM[20] and attention are typical mechanisms to visualize the regions of interest used in many fields, and there have been several reports of their application to pathological tissues[21–24]. These models often provide a heatmap, highlighting the areas that influenced the outputs, or extract representative areas for explainability. Among other advantages of such approaches are that it is easy to generalize, and the output is not restricted by existing cognitive frameworks, such as cancer cell, mitosis, and necrosis, etc. At the same time, there are significant gaps in outputs highlighted by AI-generated heatmaps and the traditional pathological approach which is the intuitive process to find out diagnostic clue in the tissue.

Here, we present a new strategy, MIXTURE (huMan-In-the-loop eXplainable artificial intelligence Through the Use of REcurrent training), to easily extract microscopic findings recognized by expert pathologists assisted by deep learning, using the histopathology of interstitial pneumonia as an example. We also show that these extracted findings can be used for practical tasks such as predicting diagnosis and analyzing prognostic factors. In this way, we are able to take advantage of computational pathology to perform quantitative studies based on well-documented pathological concepts rather than the fully automated heatmap, which leaves room for interpretation.

## 2. Materials and Methods

### 2.1. Study cohort

This is a retrospective study using a series of consulted cases (2009–2020) from a single institute. Ethical approval of this study was granted by the Ethics Committee of Nagasaki University Hospital (protocol 19012107). Three non-overlapping datasets were created from these cases, including two pretraining sets and one utility set (Figure 1). The patient characteristics in each cohort are shown in Table 1.

**Figure 1:**
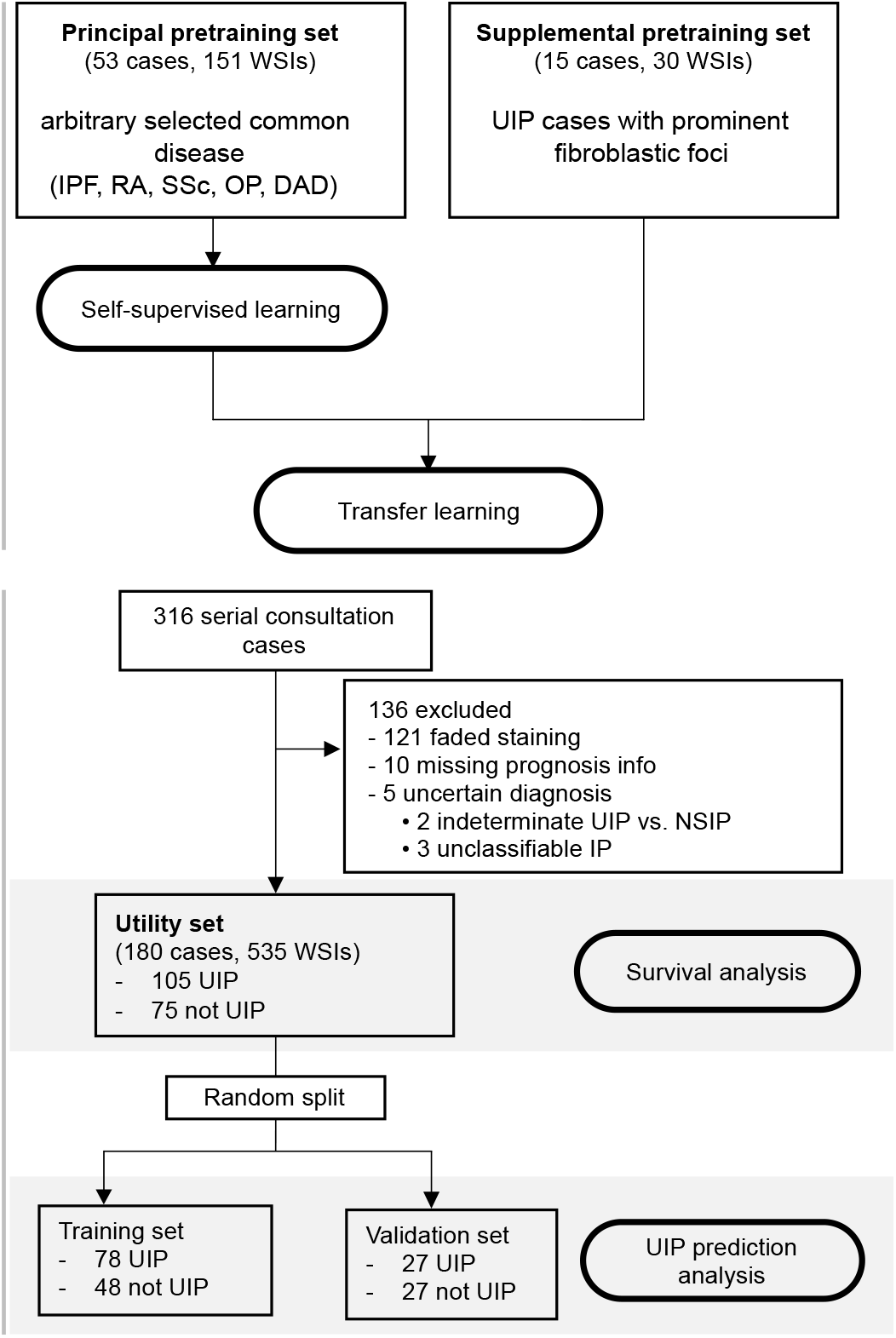
Flow diagram of the study.

**Table 1:** Patient characteristics of each cohort.

|  | Pretraining set |  | Utility set |  |
| --- | --- | --- | --- | --- |
|  | Principal<br>(n = 53) | Supplemental<br>(n = 15) | Training<br>(n = 126) | Validation<br>(n = 54) |
| Age (SD) | 59.57 (11.91) | 66.2 (7.55) | 63.27 (7.15) | 60.30 (11.18) |
| Sex |  |  |  |  |
| Male (%) | 31 (58.5) | 14 (93.3) | 74 (58.7) | 34 (55.6) |
| Female (%) | 22 (41.5) | 1 (6.7) | 52 (41.3) | 20 (37.0) |
| Sampling modality |  |  |  |  |
| SLB | 49 | 15 | 126 | 54 |
| TBLC | 3 | 0 | 0 | 0 |
| TBLB | 1 | 0 | 0 | 0 |
| UIP cases, n (%) | n/a | n/a | 78 (61.9) | 27 (50) |
| Follow up time, days (SD) | n/a | n/a | 1430.9 (469.1) | 1267.9 (461.7) |
| Event, death (%) | n/a | n/a | 29 (23.2%) | 12 (23.1%) |

SD, standard deviation; SLB, surgical lung biopsy; TBLC, transbronchial lung cryobiopsy; TBLB, transbronchial lung biopsy; UIP, usual interstitial pneumonia; n/a, not applicable

The principal pretraining set was a cohort established for the purpose of building a model to classify tiles; cases were arbitrarily selected from those sampled between 2015 and 2020 with the aim of covering a variety of histological patterns important in diagnosis and differential diagnosis of interstitial pneumonia. This set consisted of 53 cases (151 WSIs), mainly from the five most frequent diseases belonging to the interstitial pneumonia family (IPF/UIP, rheumatoid arthritis, systemic sclerosis, diffuse alveolar damage, pleuroparenchymal fibroelastosis, organizing pneumonia, and sarcoidosis).

The supplemental pretraining set (15 cases, 30 WSIs) was a cohort selected to extract rare but important histopathological findings such as fibroblastic foci[15, 25–28]. This set consisted of surgical lung biopsy specimens consulted between 2015 and 2020 in which fibroblastic foci were prominent.

The utility set consisted of 180 consecutive surgical lung biopsy cases (535 slides) sampled between 2009 and 2014 for which follow-up data was available. WSIs that were not suitable for analysis, such as those with faded staining, were excluded. All cases were diagnosed by an expert pulmonary pathologist (J.F.) and thoroughly reviewed in multidisciplinary discussion with clinicians and radiologists (supervised by T. J. and Y. K. as senior experts).

### 2.2. Image preparation

Glass slides were scanned at 20x magnification into digital slides using an Aperio ScanScope CS2 digital slide scanner (Leica Biosystems, Buffalo Grove, IL).

Figure 2 shows the overview of the following procedures of MIXTURE. In the principal pretraining set WSIs were tiled into non-overlapping 280 × 280 pixel images at magnifications of 2.5x, 5x, and 20x, respectively. Three different magnifications were studied because they provide access to different and sometimes non-overlapping morphological findings (described below in a section about labelling/clustering) having important diagnostic significance for evaluation of interstitial pneumonia. Background was defined as pixels with all values above 220 in the 24-bit RGB color space, and tiles with more than 90% of this coverage were excluded. If more than 300 tiles were obtained from a single slide, 300 tiles were randomly selected. Finally, we collected 36,978 tiles for 2.5x magnification, 44,066 tiles for 5x magnification, and 45,300 tiles for 20x magnifications.

**Figure 2:**
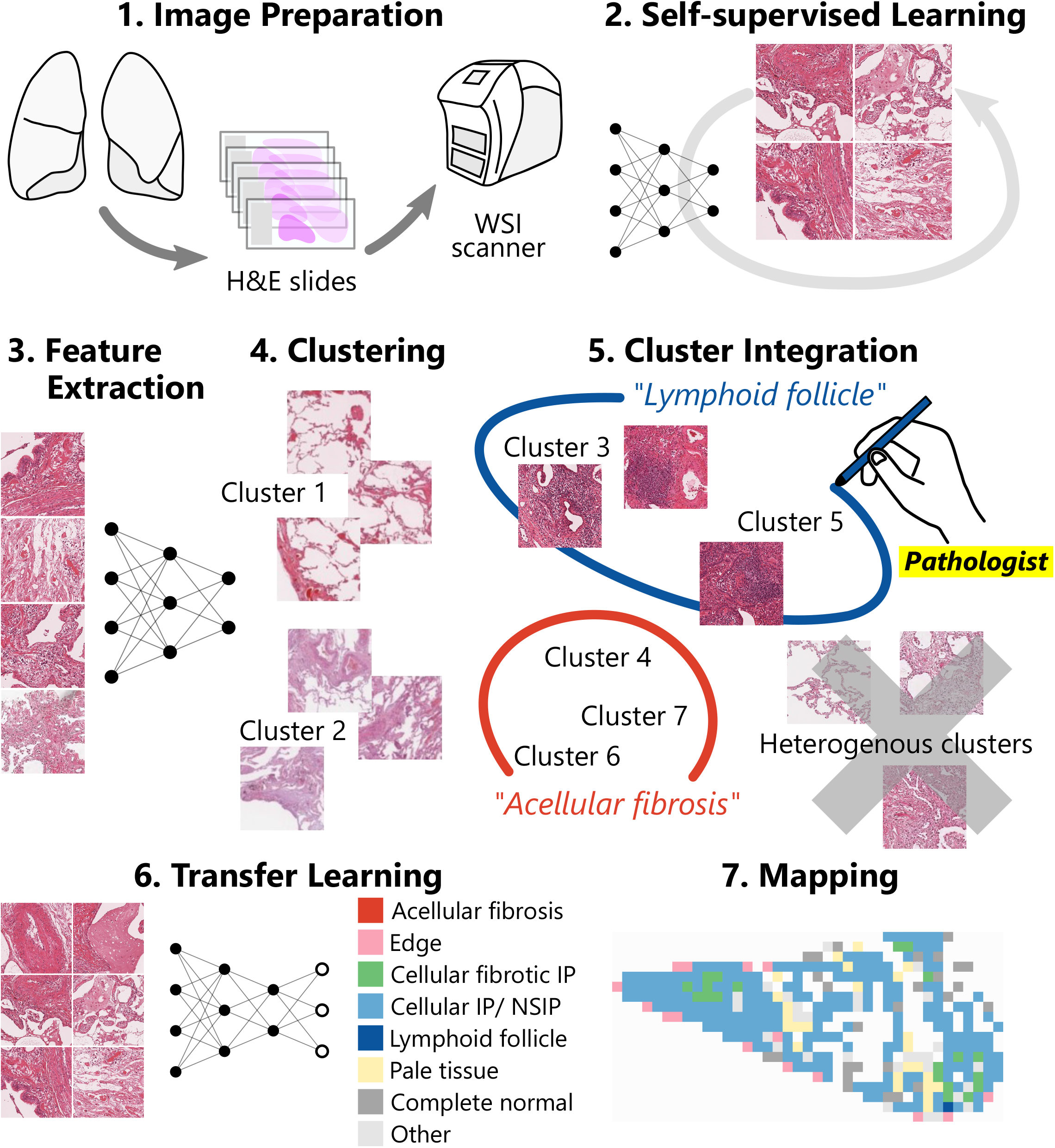
Pipeline overview of MIXTURE. For each magnification, elemental feature extractors (ElEx) were trained using self-supervised learning. This feature extractor consists of a ResNet18 CNN which outputs features consisting of 128 vectors. The extracted features were clustered throughout the principal pretraining set. The pathologists viewed a montage of each cluster tiles and reclassified them into pathologically meaningful findings. Finally, the reclassified findings were used as labels of training data for the transfer learning of feature extractor to obtain a classifier to classify the findings from the tiles.

In the supplemental pretraining set, WSIs were tiled into 280 × 280 pixel images with 50% overlap at 20x magnification. Tiles over 70% background were excluded; all images were used, regardless of the number of tiles generated from a single WSI.

In the utility set, WSIs were tiled into non-overlapping 224 × 224 pixel images. Tiles over 70% background were excluded; all images were used, regardless of the number of tiles generated from a single WSI.

### 2.3. Development of elementary feature extractor (ElEx) by selfsupervised learning

We first used the tiles from the principal pretraining cohort to create an elementary feature extractor (ElEx), which will be the basis for clustering similar tiles and for later transfer learning.

We trained a CNN (ResNet18) that outputs features consisting of 128 vectors by self-supervised learning (MoCo [29]) for each of three magnifications (2.5x, 5x, 20x). The original algorithm uses multiple GPUs, but due to the limitations of our computational resources, we modified a single GPU version[30] available for Google Colab[31]. The number of negative keys (moco-k) was set to 4096, moco momentum of updating key encoder (moco-m) was set to 0.99, and softmax temperature (moco-t) was set to 0.1.

During training, each image was randomly flipped and rotated between -20° and 20°, and central 224 × 224 pixels were cropped to make it compatible with the original dimensions of ResNet18. We used Adam as the optimizer with a global learning rate of 0.0001.

### 2.4. Clustering of tiles

The tiles in the principal pretraining set were converted into feature vectors comprised of 128 values by the ElEx we developed in the previous step. To aggregate similar images, these feature vectors were clustered using the K-means algorithm for each magnification. To provide a comprehensive view of the pathological findings that characterize each cluster, a montage (Figure S1) was created by randomly selecting 120 tiles from each cluster. We tested various numbers of clusters: 5, 8, 10, 30, 50, 80, 100, and 120 clusters. A small number of clusters tended to contain multiple findings within a single cluster, while a large number of clusters tended to contain the same findings in multiple clusters. In other words, using too few clusters was overly broad, and using too many clusters became redundant. The pathologist used the generated montage as a reference to determine the findings to be classified by each magnification and selected the optimal number of clusters.

### 2.5. Cluster integration and transfer learning

Two pathologists (J.F. and W.U.) reviewed the montages and grouped clusters characterized by pathologically synonymous findings into separate classes. The morphological findings we categorized were the following (Figure S2): for 2.5x magnification, acellular fibrosis, cellular fibrosis, near normal, and other; for 5x magnification, acellular fibrosis, edge, cellular and fibrotic IP, cellular interstitial pneumonia/NSIP, lymphoid follicle, complete normal, and other; for 20x magnification, dense fibrosis, elastosis, fibroblastic foci, fat, mucin, bronchiolar epithelium, lymphocyte aggregation, and other. The “edge” in 5x means the sharp structural contrast to airspace and the “pale” in 5x includes tiles with faded staining or structures refractory to H&E staining (e.g. elastic fibers). In order to comprehensively investigate the relationship between findings, morphologically recognizable findings were adopted as independent findings, even when their significance was unknown. Clusters that did not fit into any of the findings or were difficult to explain as morphological findings were grouped into a single class, “other”. Clusters that characterized more than one morphological finding (e.g., a cluster which had both “acellular fibrosis” and “cellular fibrotic IP”) were excluded. Thus, labeled data was constructed with the aid of ElEx clustering. We term this process “cluster integration”, meaning the merging of synonymous clusters together and the cleaning up of cluster data by pathologists.

Although the 20x resolution tiles could be labeled “dense fibrosis”, “bronchiolar epithelium”, or “lymphocyte aggregation” by this procedure, clusters consisting purely of fibroblastic foci, one of the most important findings, could not be obtained, even when the number of clusters was quite large. In order to collect these important findings, we clustered the tiles of the supplemental pretraining set, which was enriched with a large number of fibroblastic foci, by case. In this way, we obtained clusters of purer findings, and we added these to the labeled data. In addition, we checked the labeled data only at 20x resolution and manually corrected the mislabeled data.

We added a fully connected layer on top of the ElEx and created CNN classifiers of morphological findings by transfer learning, in which the integrated classes were used as labels of training data (Figure 3). The loss function was defined as the cross entropy between predicted probability and the true class labels, and we used Adam optimization with a learning rate of 0.0001. In this step, instead of only optimizing the weights of the fully connected layer, we also optimized the parameters of previous layers, including all convolution filters of each layer.

**Figure 3:**
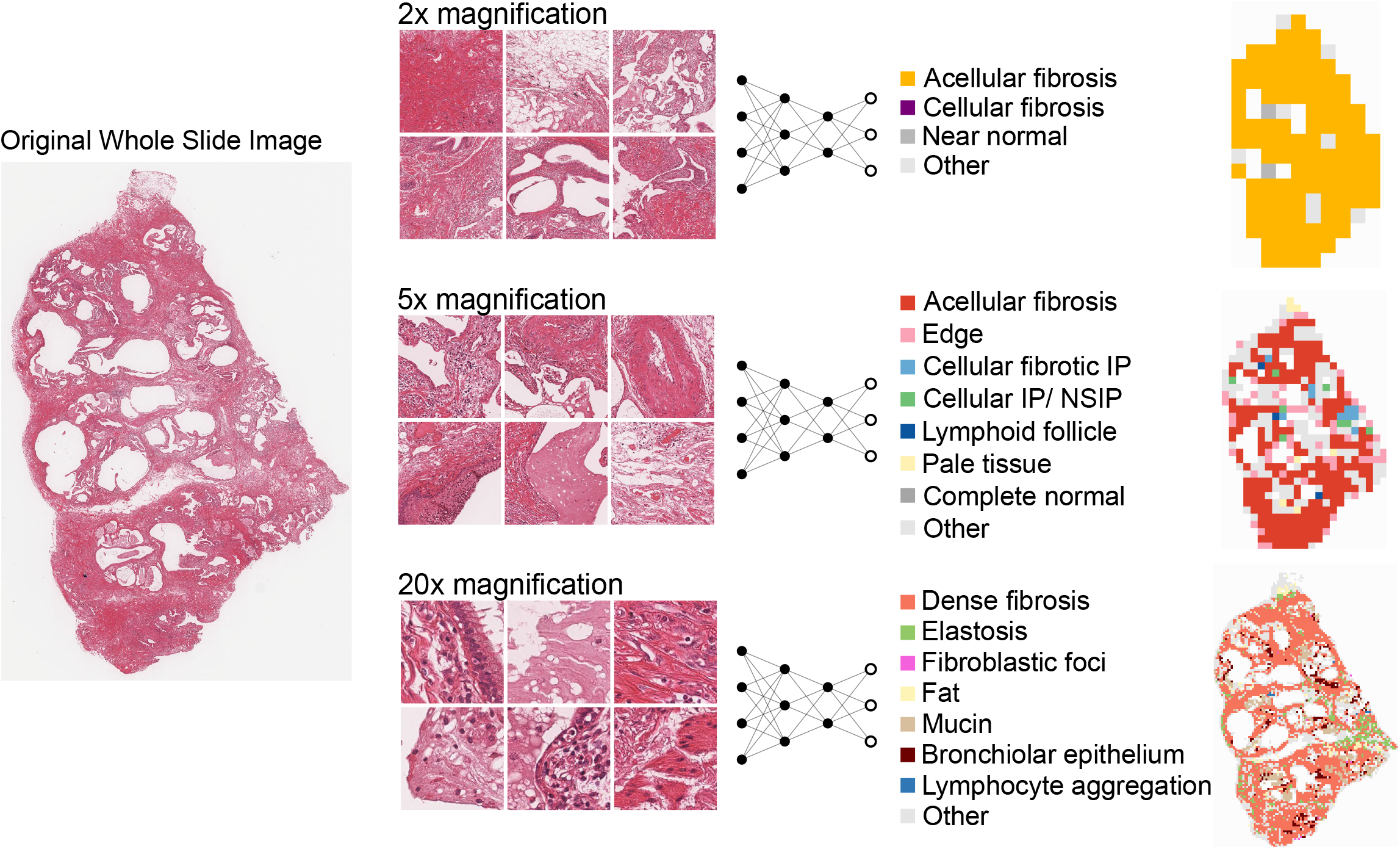
Identification of findings at each magnification. From the whole slide image, tiles were created at 2.5x, 5x, and 20x magnifications. For each magnification, a CNN classifier was constructed to classify each tile into multiple findings. Based on the classification, maps that can be compared with WSI were synthesized.

### 2.6. Tile classification and mapping of findings on WSIs

The tiles obtained from the utility set were classified using the CNN classifier created in the previous step. The results were mapped and compared with the original WSIs by two pathologists (J.F. and W.U.). In order to use the classifications for subsequent analysis, the results obtained for each case were aggregated, and the number of tiles predicted as each finding was totaled. When there was more than one WSI in a case, all tiles collected were added together. Considering the possibility that the size of the normal lung area in a surgical specimen may vary depending on the sampling procedure, tiles classified as “complete normal” were excluded at 5x magnification, and the frequency of other findings was calculated. (Note that many tiles originating from normal lungs have already been excluded because tiles containing more than 70% background were excluded at the time of the image preprocessing.)

### 2.7. UIP prediction

The UIP pattern is known as a histological pattern which characterizes IPF, furthermore, it indicates a progressive clinical course and poor prognosis with short overall survival in other interstitial lung diseases[32, 33]. Based on the well-known fact that UIP pattern is a key predictor of adverse outcome in IPF[16, 34, 35], our cases were dichotomized into UIP and non-UIP groups. We considered that this binary classification coupled with an overall survival as an endpoint may reliably estimate the performance of our AI model from a clinical point of view. We defined UIP as cases diagnosed with “definite UIP” or “probable UIP” in the pathology report and non-UIP as all other cases according to the international 2011 guidelines[36]. The 180 patients in the utility set were randomly assigned into a training set of 126 cases and a validation set of 54 cases. UIP prevalence was balanced between the training and validation set.

We developed both random forest and support vector machine models to predict UIP/non-UIP based on the frequency of each finding. In the validation set, these models were applied to predict UIP/non-UIP, and the area under the receiver operating characteristic curve was calculated to evaluate the performance for actual diagnosis. We tested whether the diagnosis of UIP predicted by the proposed model could predict the overall survival by using the log-rank test.

### 2.8. Comparison of non-integrated model and MIXTURE

To assess the effects of cluster integration by pathologists and subsequent transfer learning, we created a model without these steps (non-integrated model). The tiles from the principal pretraining set were divided into 4, 8, 10, 20, 50, and 80 clusters based on the feature vector generated by ElEx. Tiles derived from the utility sets were also converted into feature vector and the nearest cluster was predicted referring the centroid of each cluster in the previous step. As in the original models, maps of findings associated with WSIs were created, and the frequency of each finding at each magnification was calculated.

We also developed both random forest and support vector machine models to predict pathological diagnosis of UIP using this frequency of the clusters. We evaluated how the receiver operating characteristic (ROC) curve and its area under the curve (AUC) were affected when we used non-integrated model instead of MIXTURE based proposed model. The statistical significance between the AUCs from the different models was estimated by 5,000 iterations of the bootstrap method.

### 2.9. Analysis of factors associated with survival

We examined the histological risk factors for short overall survival using all cases in the whole utility set with the Cox proportional hazard model. Similarly, the histological risk factors were also estimated in the subgroups, which pathologist diagnosed as UIP and non-UIP.

### 2.10. Environment

All of the analysis in this study was executed on a Ubuntu 20.04 Linux system with a single GPU (NVIDIA RTX 3090). WSIs was tiled using the OpenSlide[37] library. Deep learning was performed using Pytorch[38], python library version 1.7.1 with CUDA 11 and cuDNN 8.0.2. K-means clustering was performed in scikit-learn version 0.24.0. The analysis for the extracted morphological findings was performed in R version 3.6.3[39]. We used the randomForest 4.6.14 package for the random forest algorithm, the pROC[40] 1.6.12 package for ROC analysis, and the survival 3.1.8 package for survival analysis.

## 3. Results

### 3.1. Tile classification and visualization

By the observation of clustered images by pathologists, the numbers of clustering were set as 30, 80, and 80 for 2.5x, 5x and 20x magnification, respectively. Using the CNN classifier we built by transfer learning, all tiles were categorized into several findings. Figure 4 shows the original WSIs and the finding maps at magnifications of 2.5x, 5x, and 20x. Additional examples are given in the Figure S3.

**Figure 4:**
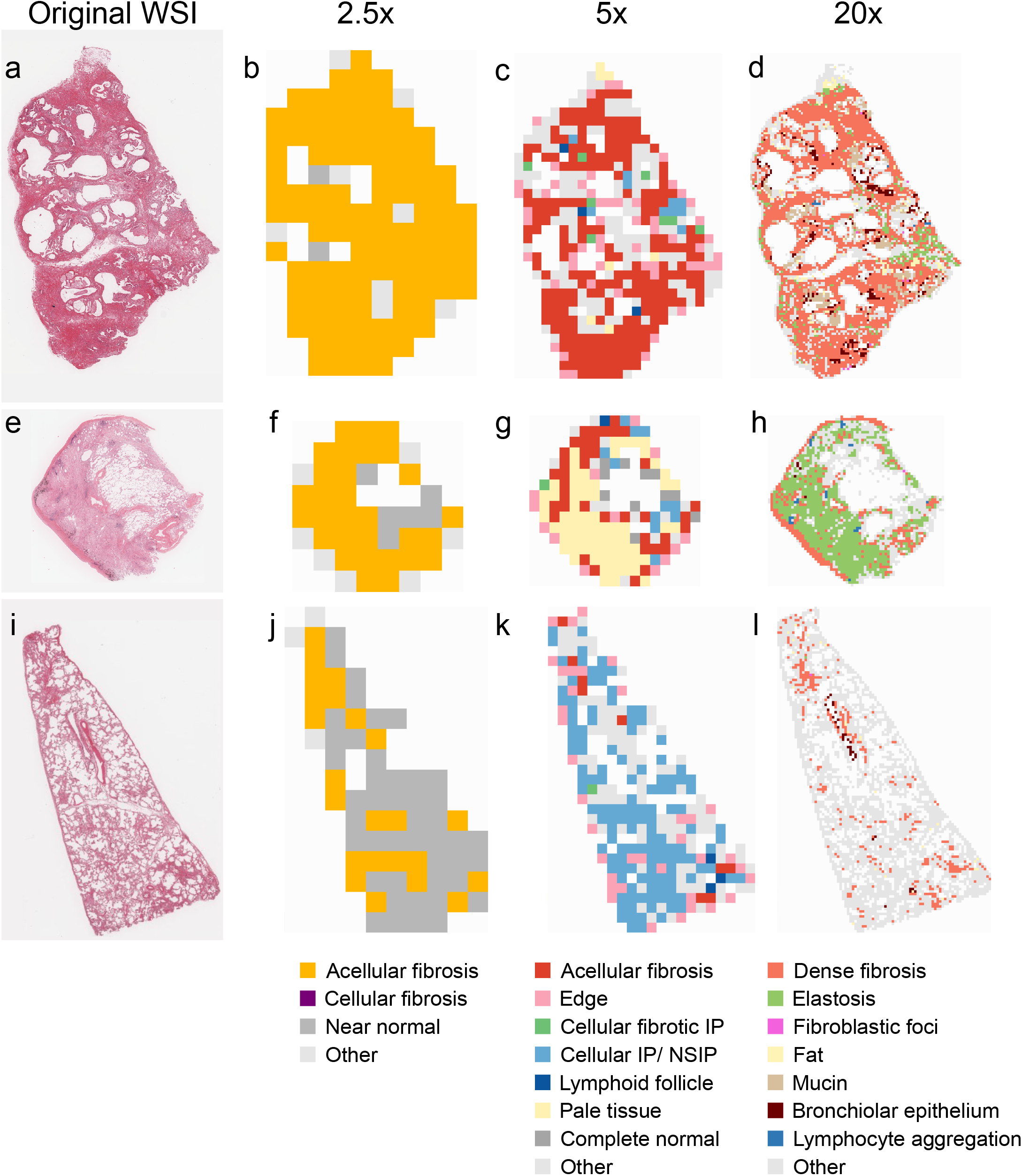
Classification of findings in the representative entities. a-d. UIP/IPF case. The entire specimen consists of dense fibrosis with minimal inflammatory cell infiltration, and is highlighted in yellow, red, and orange at 2.5x, 5x, and 20x magnification, respectively. Elastosis and bronchial metaplasia at the margins of the specimen are appropriately highlighted at 20x. e-h. Idiopathic pleuroparenchymal fibroelastosis (PPFE) case. A subpleural band of elastosis is clearly visualized by the 20x feature extractor. The same finding is recognized as “pale” tissue in 5x. i-l. A case of NSIP in systemic sclerosis. The pathology shows cellular and fibrotic NSIP, which is clearly differentiated from UIP lesions by blue highlighting on 5x feature extractor. a b

The histological findings observed in characteristic tissue patterns such as UIP and NSIP were displayed with good contrast, and a side-by-side comparison between WSIs and the maps were made to confirm that these findings were appropriately detected.

### 3.2. UIP prediction by MIXTURE

We developed a random forest model to predict the diagnosis of UIP by pathologists using the findings extracted at 5x magnification, and the model was able to predict the diagnosis with AUC 0.90 in the validation cohort (Table 2). Similarly, the models based on the findings of 20x magnification, and the combination of 20x with other studied magnifications also predicted the diagnosis of UIP with high accuracy. The ROC curves are shown in Figure 5a and Figure 5b, and the relationship between the score of the random forest regressor and the actual pathology diagnosis is shown in Figure S4. The most important findings in the random forest model were cellular interstitial pneumonia/NSIP and acellular fibrosis (Table 3). Feature importance in the models on other magnifications are shown in Table S1-S4. There were no significant differences in performance between models using only findings extracted at 5x magnification, findings extracted at 20x magnification, or a combination of these findings from different magnifications. However, it was difficult to predict UIP using only the findings extracted at 2.5x magnification. When the threshold for judging UIP was set to 0.5 for the output of the random forest regressor in 5x model, cases predicted to be UIP had a poorer prognosis than those predicted to be non-UIP (Figure 5d): five-year overall survival was 55.4% in cases predicted as UIP whereas 95.2% in cases predicted as non-UIP.

**Table 2:** AUC for each model.

| Table 2: AUC for each model |  |  |
| --- | --- | --- |
|  | AUC | 95% CI |
| Proposed model |  |  |
| 2.5x | 0.68 | 0.54 – 0.83 |
| 5x | 0.90 | 0.81 – 0.99 |
| 20x | 0.90 | 0.81 – 0.99 |
| 2.5x + 5x | 0.88 | 0.78 – 0.98 |
| 5x + 20x | 0.92 | 0.85 – 1.00 |
| 2.5x + 20x | 0.89 | 0.80 – 0.98 |
| 2.5x + 5x + 20x | 0.92 | 0.84 – 1.00 |
| Non-integrated model (5x) |  |  |
| k = 4 | 0.52 | 0.37 – 0.68 |
| k = 8 | 0.65 | 0.50 – 0.81 |
| k = 10 | 0.49 | 0.33 – 0.65 |
| k = 20 | 0.47 | 0.31 – 0.63 |
| k = 30 | 0.61 | 0.46 – 0.76 |
| k = 50 | 0.56 | 0.40 – 0.72 |
| k = 80 | 0.52 | 0.36 – 0.68 |

AUC, area under the receiver operator characteristic curve; CI, confidence interval; k, number of clusters

**Figure 5:**
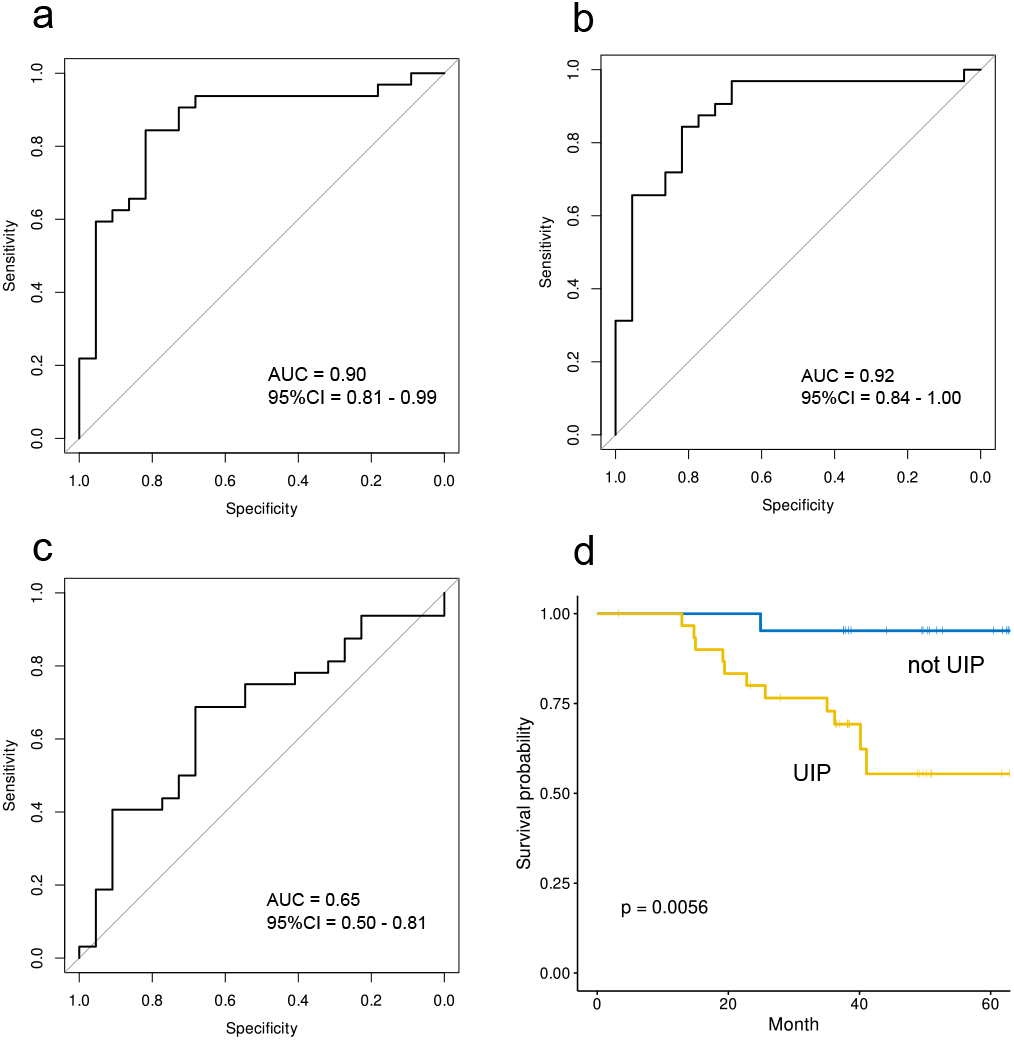
Receiver operating characteristic curves for our model’s classifications on the independent validation set. a. ROC curve when the findings obtained at 5x are used to predict the presence of UIP. b. ROC curve when all findings obtained at 2.5x, 5x, and 20x are used. c. ROC curve without pathologist integration of findings and subsequent transfer learning (non-integrated model). The case with the best AUC (k = 8) is presented. d. Model created using tiles extracted at 5x magnification. Cases predicted as UIP had a significantly worse prognosis than those predicted as non-UIP.

**Table 3:** Feature importance (node purity) of each finding in 5x model with random forest algorithm.

| Findings | Importance |
| --- | --- |
| Acellular fibrosis | 4.80 |
| Cellular and fibrotic IP | 3.80 |
| Cellular IP/NSIP | 7.54 |
| Lymphoid follicle | 2.82 |
| Edge | 4.68 |
| Pale | 3.24 |

IP, interstitial pneumonia; NSIP, non-specific interstitial pneumonia

Instead of the random forest, support vector machines were used to predict the diagnosis of UIP. The results are shown in Table S5. As in the case of the random forest, the diagnosis of UIP could be predicted with high accuracy.

### 3.3. UIP prediction by non-integrated model

In order to test the effectiveness of the pathologist’s integration of the clusters and subsequent transfer learning, we developed another model without cluster integration by a human pathologist (non-integrated model) and the performance of UIP prediction was compared. The original WSI and the maps of the tile classifications were compared, and pathologists (J.F. and W.U.) confirmed that tiles characterized by similar pathological findings were categorized in the same cluster. In addition, we examined whether UIPs could be predicted from the distribution of the predicted clusters. The number of clusters we evaluated ranged between 4 and 80; we found that the best results were obtained when assorting into 8 clusters on 5x magnification, but the AUC only reached 0.65 (Table 2). ROC of non-integrated model is shown in Figure 5c. There was a significant difference (p = 0.0002) in performance compared to the MIXTURE-based model.

Similar results were obtained when we used support vector machine instead of random forest (Table S5). Eventually, non-integrated model could not achieve high accuracy in UIP diagnosis irrespective of the type of prediction algorithm (random forest or support vector machine) and number of clusters.

### 3.4. Factors associated with patient survival

Next, to identify histological risk factors for survival, all cases in the utility cohort were examined by the Cox proportional hazards model. Since we extracted similar findings at different magnifications, we observed pairs of findings that were highly correlated in frequency within a case (Figure S5). To avoid multicollinearity, variables with high correlation, such as acellular fibrosis (2.5x), near normal (2.5x), acellular fibrosis (5x), and lymphoid follicle (20x) were excluded prior to analysis. The independent prognostic factors identified in this analysis were fibroblastic foci, dense fibrosis, elastosis, and dense lymphocyte aggregation (Table 4). In a subgroup analysis of cases diagnosed with UIP by pathologists, only fibroblastic foci were a poor prognostic factor (Table S6). Interestingly, lymphocyte aggregation was identified as a poor prognostic factor in patients diagnosed as non-UIP by pathologists (Table S7), which is not usually well acknowledged.

**Table 4:** Analysis of prognostic factors by Cox proportional hazards model.

|  | Hazard ratio | 95% CI | p value |
| --- | --- | --- | --- |
| Cellular fibrosis | 0.83 | 0.57 – 1.22 | n.s. |
| Cellular IP/ NSIP | 0.84 | 0.48 – 1.47 | n.s. |
| Edge | 1.10 | 0.78 – 1.55 | n.s. |
| Dense fibrosis | 1.57 | 1.04 – 2.40 | 0.034 |
| Fibroblastic focus | 1.47 | 1.11 – 1.96 | 0.008 |
| Elastosis | 1.48 | 1.02 – 2.15 | 0.040 |
| Fat | 1.15 | 0.85 – 1.57 | n.s. |
| Lymphocyte aggregation | 1.35 | 1.03 – 1.77 | 0.030 |
| Mucin | 1.17 | 0.81 – 1.69 | n.s. |
| Bronchiolar epithelium | 0.74 | 0.50 – 1.10 | n.s. |

IP, interstitial pneumonia; NSIP, non-specific interstitial pneumonia; n.s., non-significant

## 4. Discussion

In this study, we proposed a method, MIXTURE, to build a deep learning model without laborious direct annotations and showed this model working effectively in the pathology field. In this method, the encoder specialized in pathological images was developed by self-supervised learning and used to cluster the tiles which have similar morphological findings. Pathologists integrated the morphologically synonymous clusters into several classes, which were used as training data for subsequent transfer learning. The model illustrates the amount and the distribution of each morphological finding compared with the original WSI, which was utilized to build an explainable AI to predict UIP diagnosis for subsequent analysis.

The unique point of this method is that the images that are clustered based on similarity are further integrated by experts and used as training data. There are three advantages to using this method. The first is that it leaves room for the expert’s judgment in model creation. In reality, K-means clustering alone does not always form pathologically meaningful clusters, and may form clusters based on non-essential characteristics such as differences in staining or specimen condition. It is considered that the integration of the clusters may extenuate these non-essential differences. The method to integrate the clusters depends on the insights of the experts, which may affect the final model. In fact, the result that the UIP could not be predicted without the integration process suggests that the performance of the final model could be greatly affected by this step. The second advantage is that clustering reduces the huge cost of labeling for each tile. Tile labeling requires expertise and needs to be optimized for each application. Thus, annotation of pathological tissues costs a lot of time and money. However, there is a chronic shortage of pathologists[41, 42], making it almost impossible to obtain a large number of annotations in reality, i.e. in clinical settings. The third advantage is that clustering over the entire dataset makes it easier to maintain the consistency of the training data. Many pathological findings are essentially continuous and change without a distinct boundary, especially in benign diseases such as interstitial pneumonia, and judgments are often not consistent between evaluators[15–17]. Therefore, elaborate annotation of such findings is difficult, and even if it were possible, there is concern that these differences between the individuals and the timing of the annotations will result in inconsistent training data.

There are several points that should be considered concerning clustering. The histological findings that characterize the clusters depend on the size and resolution of each tile. Therefore, we need to set the appropriate magnification and tile size according to the required findings. In addition, there are findings such as adipocytes and loose stromal tissue that are easily recognized by pathologists but tend to be classified into the same cluster. In this case, manual labeling was more effective to create training data. This was often true for well-defined findings that could be identified with high magnification. Even when the tiles were manually labeled, clustering improved the efficiency of the task.

The proposed approach does not adopt an end-to-end learning structure, which is common in state-of-the-art research[22, 43]. End-to-end learning directly outputs the result, bypassing the feature extraction steps. The performance of the system is generally high because it is relatively free from potential human cognitive biases, but the decision-making mechanism is a black box. Although recent models are designed to highlight the areas that contribute to the output[22, 44], it is still necessary to reinterpret the output from an expert perspective. In reality, the cases in which pathologically useful findings have been discovered from these explanations are quite limited.

In contrast to these approaches, we designed a model that outputs findings. In conventional pathological observational studies, these findings are implicitly identified by experienced pathologists. This process can be naturally replaced by deep learning. Additionally, the output is easily interpretable by pathologists without any AI background for use in subsequent analysis, and it is uniquely compatible with conventional pathological knowledge. This explainability is essential in introducing in clinical setting. Our model can be integrated in the daily practice and supports pathologists by highlighting the important findings or by suggesting potential diagnoses.

When searching for certain target findings, there is a common need to quantitatively analyze histological findings. Our model seeks to serve that purpose. At present, we do not take into account the spatial relationship of each finding, but once this is implemented, more detailed analysis will be possible.

Another feature of our model is that it is composed of three independent modules. Each of them is a simple CNN that can be interpreted by itself and can be used for other tasks such as predicting treatment response. In this use case, we assigned three modules with different magnifications of 2.5x, 5x, and 20x, which simulates the actual pathological evaluation process and is intuitive for pathologists. Furthermore, if these modules are augmented with those for interpreting radiological images and genetic data instead of WSIs, it will open the door to the realization of explainable multimodal models[45], which will allow for new analytical opportunities such as interdisciplinary relationships between findings.

From a medical point of view, it is the first model known to predict the diagnosis of UIP from histopathological images. While not directly addressed in this study, other interstitial pneumonias, such as pleuroparenchymal fibroelastosis or NSIP, can be predicted in a similar way, since the characteristic spatial distributions of the findings for each disease are handled deftly by our ElEx. The random forest algorithm can estimate the importance of each finding, and our model showed that the presence of NSIP and dense fibrosis were important. This is consistent with the existing literature and actual practice[1]. In the prognostic analysis, fibroblastic foci, dense fibrosis, elastosis, and lymphocyte aggregation were identified as risk factors. Although the conclusions are controversial, some studies have mentioned the relationship between excessive fibroblastic foci and prognosis[15, 25–28]. The amount of dense fibrosis is also a diagnostic factor for UIP[1], which makes sense from a pathological point of view, and there have been reports that increased fibroelastosis is associated with poor prognosis[46]. In our data, dense inflammatory cell infiltration was identified as an independent risk factor; a similar result was obtained in the subgroup analysis of the non-UIP cohort, but it was not an independent risk factor in the UIP cohort. Related previous literature has linked interstitial mononuclear cell infiltration to respiratory function decline at 6 months in IPF patients[26]. Another group has discussed the relationship between CD3-positive T cell infiltration and poor prognosis in idiopathic interstitial pneumonia[47]. To the best of our knowledge, there are no studies that have examined the relationship between inflammatory cell infiltration and poor prognosis, especially in non-UIP patients; more studies are needed in the future.

There are some limitations in this study. First, the data used in this study were specimens collected and processed at a single institution and scanned with a single model of WSI scanner. Therefore, external validation is necessary. In addition, most of the specimens were surgical lung biopsies sampled by a relatively invasive procedure, which is currently being replaced by the less invasive transbronchial lung cryobiopsy in some institutions. Regarding the technical pipeline of MIXTURE, the findings that can be extracted are limited to those that are clustered coincidentally, so that this method is not suitable for creating training data for findings that are extremely similar or very rare. In addition, it is difficult to incorporate findings that are not recognized by the pathologist into the model. The integration of clustering strongly depends on the judgment of the pathologists. In the present study, only two pathologists discussed and made decisions, and this may be biased. We plan to validate the model by prospectively applying it to incoming cases, including those sampled by cryobiopsy. Furthermore, we see great potential for MIXTURE to be trained and tested on entities other than interstitial pneumonia, such as tumors.

In summary, we proposed an original approach to extract multiple features that can be interpreted by pathologists with minimum annotation effort by experts. The model not only effectively describes the quantity and distribution of features for different IPF entities but is also effective in explainably predicting progressive disease and quantitatively analyzing histological features. The same approach could be applied to other areas of pathology or radiology, and represents a new direction for explanatory analytical models.

## Data Availability

Data to be provided upon a reasonable request.

## Acknowledgement

This paper is based on results obtained from a project, JPNP20006, commissioned by the New Energy and Industrial Technology Development Organization (NEDO).

The authors thank Mr. Ethan Okoshi, Department of Pathology, Nagasaki University Graduate School of Biomedical Sciences, for proofreading the English manuscript.

