## Supplementary material for "MIXTURE of human expertise and deep learning—Developing an explainable model for predicting pathological diagnosis and survival in patients with interstitial lung disease": Fig. S1-S5; Table S1-S7

### Supplemental data

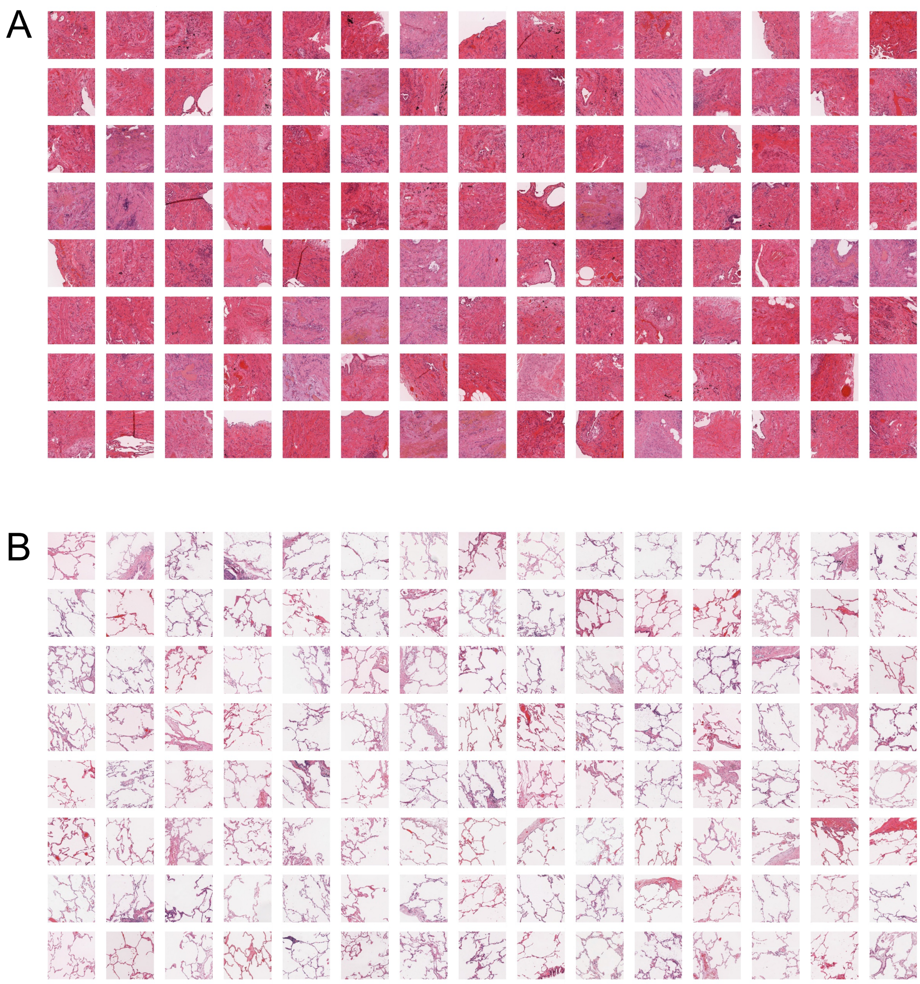


Figure. S 1: **Example of montage.** The tiles cut from each magnification were converted into feature vectors by ElEx and clustered based on the similarity. In order to demonstrate the overview of each cluster, 120 tiles were randomly selected from each cluster and a montage was created. These examples are all tiles extracted at 5x magnification from different cases; A is classified as acellular fibrosis and B as complete normal.

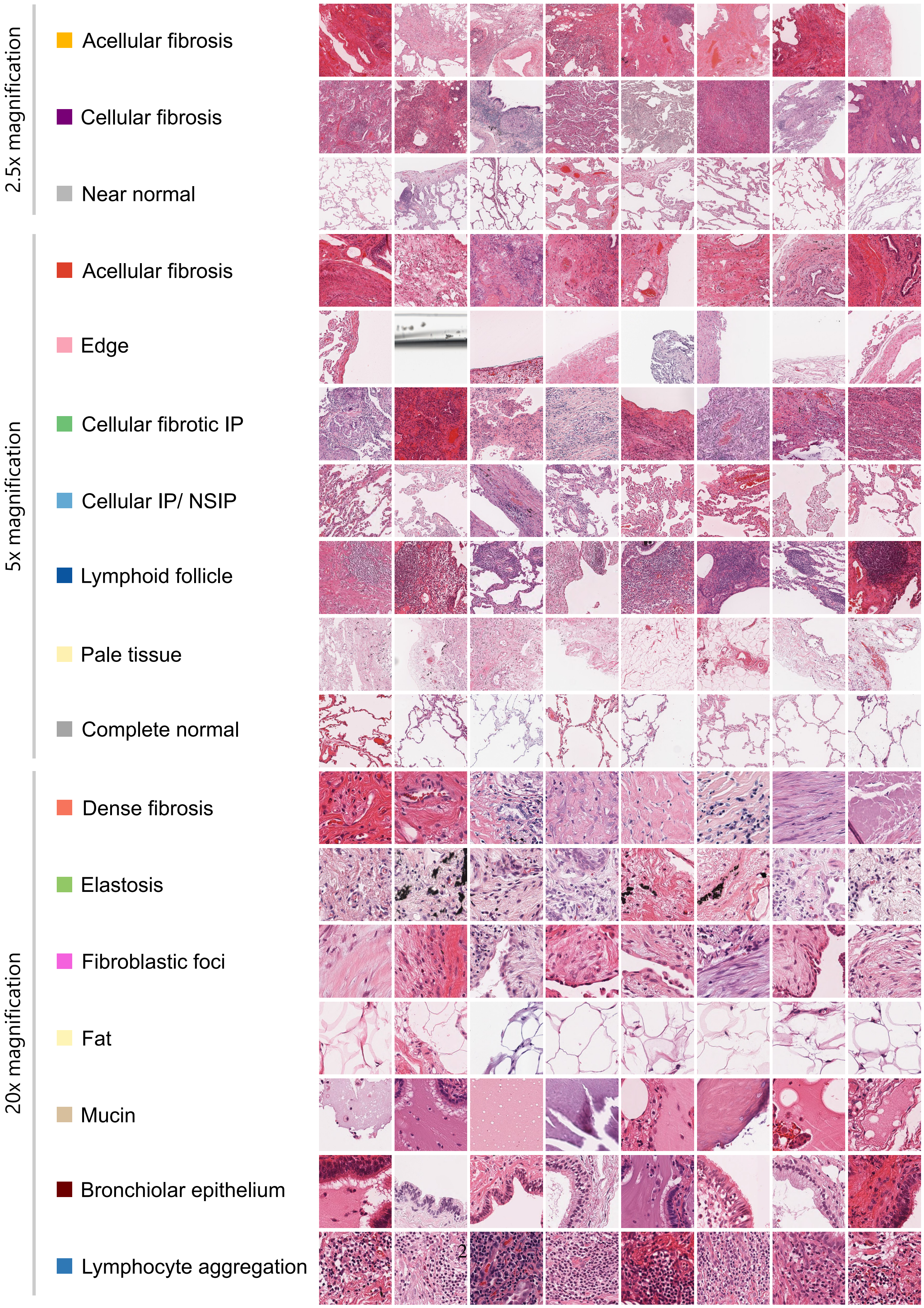


Figure. S 2: **Example of tiles for each class.**

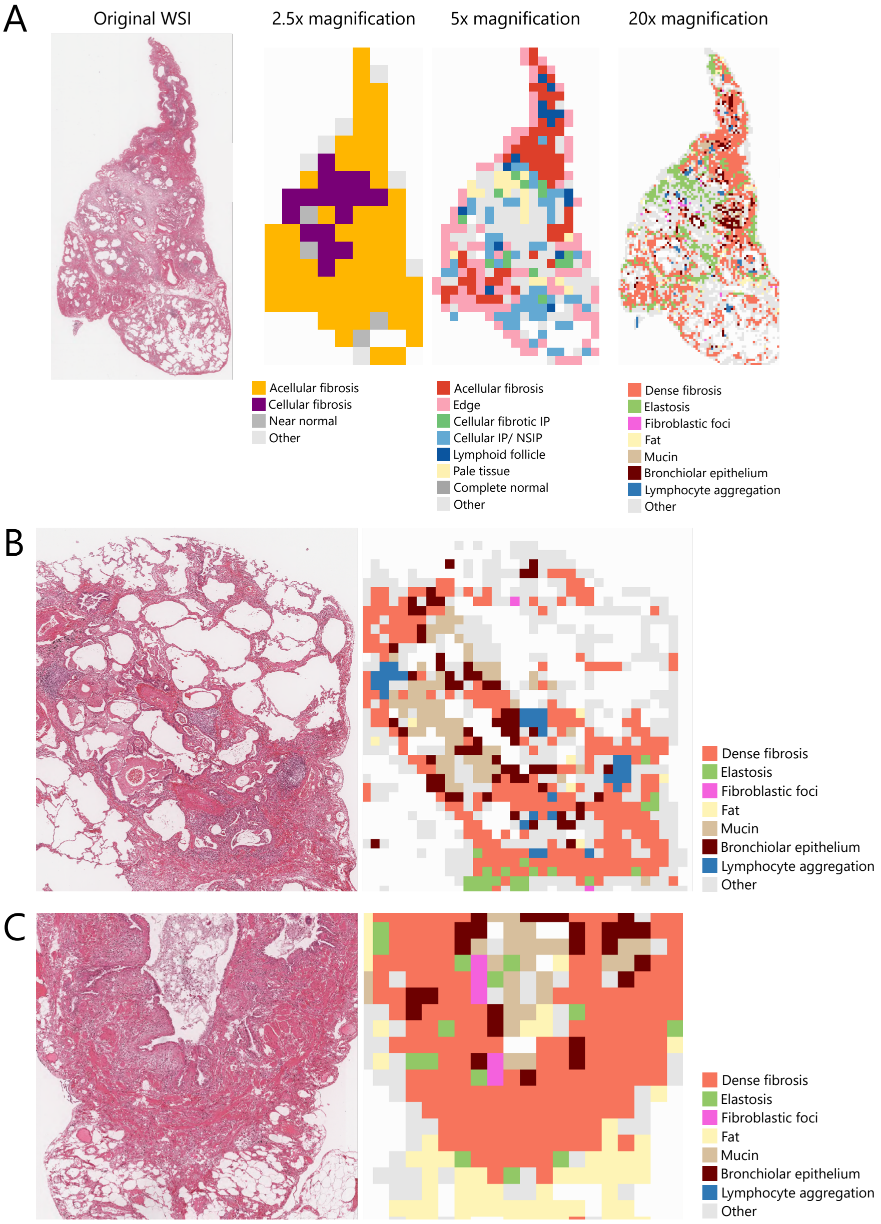


Figure. S 3: **Additional examples of feature extraction.** A: A case of interstitial pneumonia related to systemic sclerosis with a histological diagnosis of ”probable UIP”. B, C: Preview of 20x feature extractor with high magnification

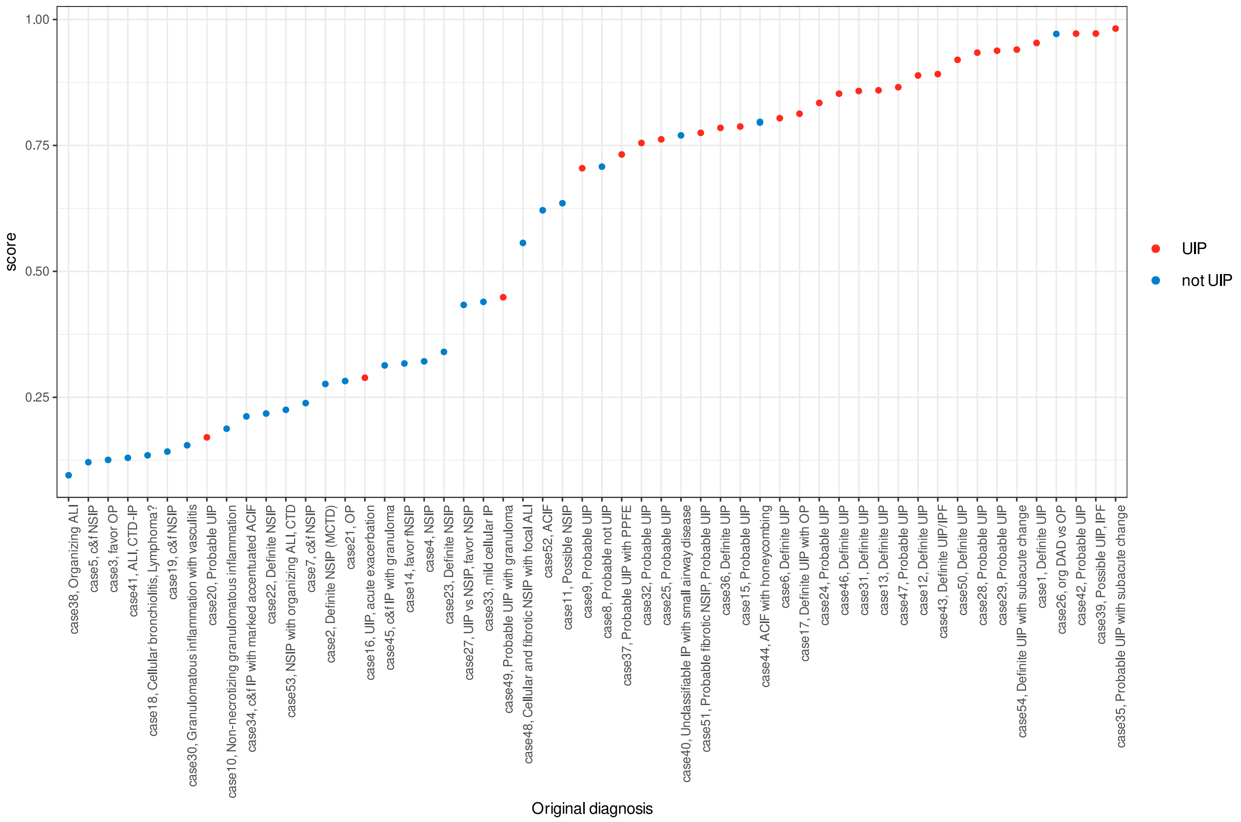


Figure. S 4: **Score of random forest regressor and the actual histopathological diagnosis.** The x-axis shows the actual pathological diagnosis for each case, and the y-axis shows the predicted UIP score by the random forest 5x model. Cases regarded as UIP from its pathology report are shown in orange, and not UIPs are shown in blue.

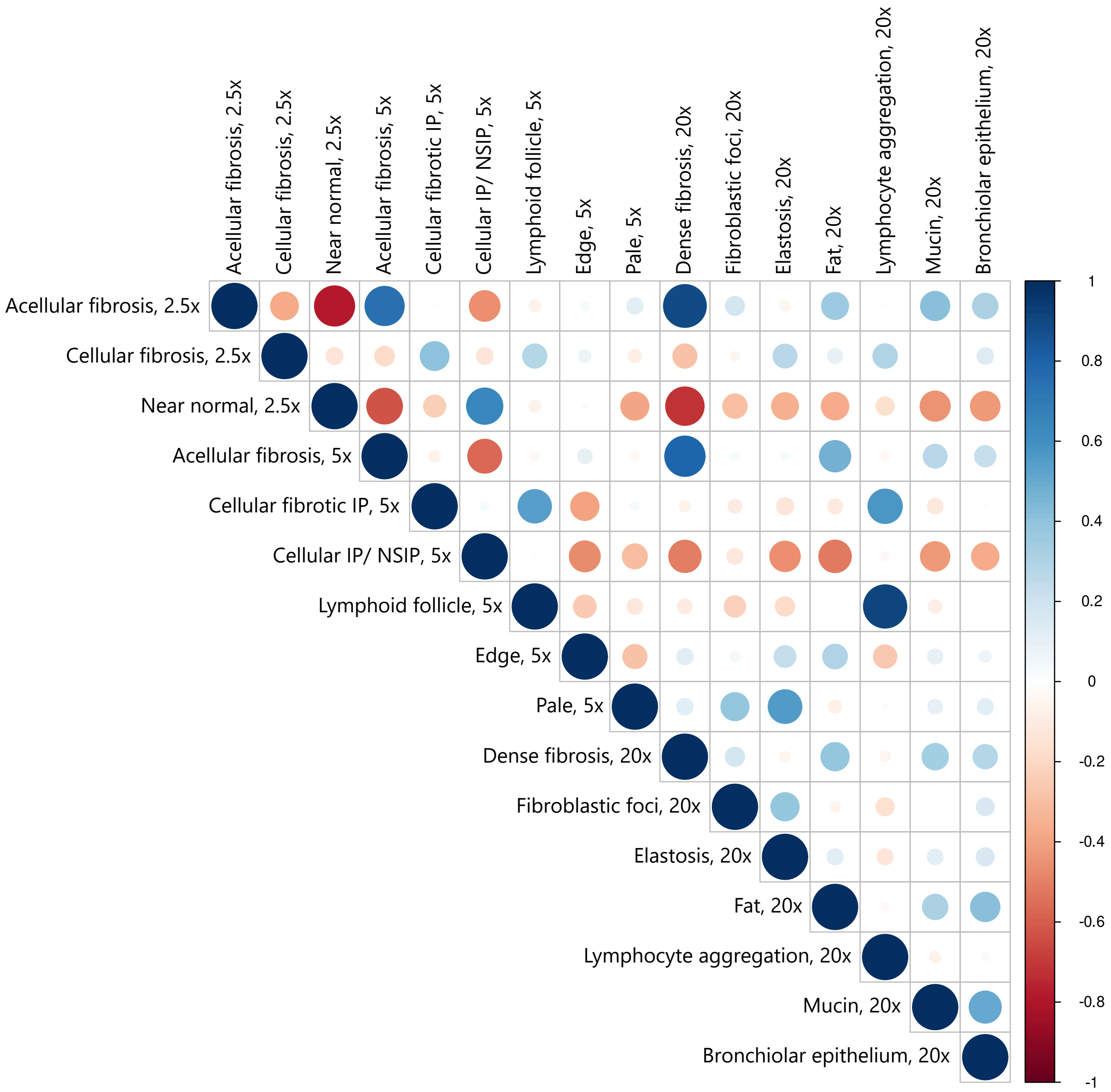


Figure. S 5: **Quantitative correlation of extracted findings.** Because similar findings were extracted for each magnification, pairs of findings with correlated occurrences were found in the same case. There is a positive correlation between acellular fibrosis extracted at 2.5x and acellular fibrosis extracted at 5x. Similarly, there is a very strong positive correlation between lymphatic follicles extracted at 5x and lymphocytes extracted at 20x.

Table. S 1: **Feature importance of 20x model**

| Findings | Importance |
|---|---|
| Dense fibrosis, 20x | 3.540 |
| Fibroblastic foci, 20x | 2.875 |
| Elastosis, 20x | 3.685 |
| Fat, 20x | 6.099 |
| Lymphocyte aggregation, 20x | 2.610 |
| Mucin, 20x | 2.765 |
| Bronchiolar epithelium, 20x | 5.218 |

Table. S 2: **Feature importance of 2.5x+5x model**

| Findings | Importance |
|---|---|
| Acellular fibrosis, 2.5x | 1.802 |
| Cellular fibrosis, 2.5x | 2.112 |
| Near Normal, 2.5x | 1.902 |
| Acellular fibrosis, 5x | 3.473 |
| Cellular and fibrotic IP, 5x | 2.724 |
| Cellular IP/ cNSIP, 5x | 7.102 |
| Lymphoid follicle, 5x | 2.234 |
| Edge, 5x | 3.598 |
| Pale, 5x | 2.335 |

IP: interstitial pneumonia, cNSIP: cellular non-specific interstitial pneumonia

Table. S 3: **Feature importance of 5x+20x model**

| Findings | Importance |
|---|---|
| Acellular fibrosis, 5x | 2.223 |
| Cellular fibrotic IP, 5x | 2.183 |
| Cellular IP/ NSIP, 5x | 5.175 |
| Lymphoid follicle, 5x | 1.336 |
| Edge, 5x | 2.124 |
| Pale, 5x | 1.644 |
| Dense fibrosis, 20x | 1.492 |
| Fibroblastic foci, 20x | 1.200 |
| Elastosis, 20x | 1.326 |
| Fat, 20x | 3.357 |
| Lymphocyte aggregation, 20x | 1.003 |
| Mucin, 20x | 1.518 |
| Bronchiolar epithelium, 20x | 3.067 |

IP: interstitial pneumonia, cNSIP: cellular non-specific interstitial pneumonia

Table. S 4: **Feature importance of 2.5x+5x+20x model.**

| Findings | Importance |
|---|---|
| Acellular fibrosis, 2.5x | 0.751 |
| Cellular fibrosis, 2.5x | 1.412 |
| Near normal, 2.5x | 0.767 |
| Acellular fibrosis, 5x | 2.030 |
| Cellular fibrotic IP, 5x | 1.850 |
| Cellular IP/ NSIP, 5x | 4.690 |
| Lymphoid follicle, 5x | 0.994 |
| Edge, 5x | 1.834 |
| Pale, 5x | 1.362 |
| Dense fibrosis, 20x | 1.356 |
| Fibroblastic foci, 20x | 0.959 |
| Elastosis, 20x | 1.282 |
| Fat, 20x | 3.415 |
| Lymphocyte aggregation, 20x | 0.899 |
| Mucous, 20x | 1.247 |
| Bronchiolar epithelium, 20x | 2.929 |

IP: interstitial pneumonia, NSIP: non-specific interstitial pneumonia

Table. S 5: **UIP prediction by support vector machine algorithm**

| | AUC | 95% CI |
|---|---|---|
| Proposed model | | |
| 2.5x | 0.87 | 0.77 - 0.97 |
| 5x | 0.89 | 0.79 - 0.97 |
| 20x | 0.9 | 0.81 - 0.98 |
| 2.5x + 5x | 0.86 | 0.76 - 0.97 |
| 5x + 20x | 0.88 | 0.79 - 0.97 |
| 2.5x + 20x | 0.89 | 0.80 - 0.98 |
| 2.5x + 5x + 20x | 0.86 | 0.77 - 0.96 |
| Non-integrated model | | |
| k=4 | 0.47 | 0.31 - 0.63 |
| k=8 | 0.55 | 0.39 - 0.71 |
| k=10 | 0.56 | 0.40 - 0.72 |
| k=20 | 0.51 | 0.35 - 0.67 |
| k=30 | 0.47 | 0.31 - 0.63 |
| k=50 | 0.61 | 0.46 - 0.77 |
| k=80 | 0.61 | 0.46 - 0.76 |

AUC: area under the receiver operator characteristic curve, CI: confidence interval

Table. S 6: **Subgroup analysis consisting of cases diagnosed as UIP by pathologists. In the Cox proportional hazards model, the amount of fibroblastic foci was confirmed to be a risk factor.**

| | HR | 95% CI | p value |
|---|---|---|---|
| Cellular fibrosis | 1.13 | 0.69 – 1.84 | n.s. |
| Cellular IP/ NSIP | 0.81 | 0.33 – 1.97 | n.s. |
| Edge | 1.22 | 0.76 – 1.98 | n.s. |
| Dense fibrosis | 1.30 | 0.80 – 2.12 | n.s. |
| Fibroblastic focus | 1.67 | 1.13 – 2.46 | 0.0010 |
| Elastosis | 1.09 | 0.65 – 1.81 | n.s. |
| Fat | 1.06 | 0.75 – 1.50 | n.s. |
| Lymphocyte aggregation | 1.27 | 0.60 – 2.71 | n.s. |
| Mucin | 1.19 | 0.80 – 1.76 | n.s. |
| Bronchiolar epithelium | 0.59 | 0.37 – 0.94 | 0.0268 |

IP: interstitial pneumonia, NSIP: non-specific interstitial pneumonia, HR: hazard ratio, CI: confidence interval, n.s.: non-significant

Table. S 7: **Subgroup analysis consisting of cases diagnosed as not UIP by pathologists. In the Cox proportional hazards model, the aggregated lymphocytes were identified as a risk factor.**

| | HR | 95% CI | p value |
|---|---|---|---|
| Cellular fibrosis | 0.19 | 0.028 – 1.26 | 0.0855 |
| Cellular IP/ NSIP | 0.83 | 0.25 – 2.76 | n.s. |
| Edge | 1.52 | 0.68 – 3.97 | n.s. |
| Dense fibrosis | 1.59 | 0.44 – 5.67 | n.s. |
| Fibroblastic focus | 1.87 | 0.69 – 5.06 | n.s. |
| Elastosis | 2.16 | 0.57 – 8.17 | n.s. |
| Fat | 0.56 | 0.29 – 10.77 | n.s. |
| Lymphocyte aggregation | 2.87 | 1.03 – 7.97 | 0.0426 |
| Mucin | 1.24 | 2.30 – 5.17 | n.s. |
| Bronchiolar epithelium | 1.02 | 0.32 – 3.25 | n.s. |

IP: interstitial pneumonia, NSIP: non-specific interstitial pneumonia, HR: hazard ratio, CI: confidence interval, , n.s.: non-significant
